# The NLP-to-Expert Gap in Chest X-ray AI

**DOI:** 10.64898/2026.02.27.26347261

**Authors:** George Fisher

## Abstract

In previous work, we achieved state-of-the-art performance on ChestX-ray14 (ROC-AUC 0.940, F1 0.821) using pretraining diversity and clinical metric optimization. Applying the same methodology to CheXpert, we received similar results when using NLP valuation and test data—but when evaluated against expert radiologist labels, performance was only 0.75-0.87 ROC-AUC. The models had learned to match the automated NLP labeling system, not to diagnose disease.

This paper documents our investigation into this failure and our suggested resolution. We identify the NLP-to-Expert generalization gap: a systematic divergence between models optimized on labels extracted from radiology reports and their agreement with board-certified radiologists. More surprisingly, we discovered that directly optimizing for small expert-labeled validation sets can be counterproductive— models with lower validation scores often generalized better to held-out expert test data.

Four findings emerged:

First, expert-labeled images for at least the validation and testing datasets, even if not for training, were vital in revealing the gap between NLP agreement and diagnostic accuracy. Without them, our models appeared excellent while failing to generalize to clinical judgment.

Second, less training is better. Short training (1-5 epochs) outperformed extended training (60+ epochs) because longer training doesn’t improve the model—it memorizes the labeler’s mistakes.

Third, ImageNet features are sufficient. Freezing the pretrained backbone and training only the classifier achieved 0.891 ROC-AUC—matching models with full fine-tuning. The rapid convergence we observed wasn’t the model learning chest X-ray features; it was the classifier calibrating to already-sufficient visual representations.

Fourth, regularization beats optimization. Label smoothing and frozen backbones—methods that prevent overfitting—outperformed direct metric optimization on small validation sets. The 200 expert-labeled validation images in CheXpert are too few to optimize directly; they are better used as a compass than a target.

With these insights, we improved from 0.823 to 0.917 ROC-AUC, exceeding Stanford’s official baseline (0.907).

## 1 Introduction

### 1.1 The Problem We Didn’t Expect

In October 2025, we submitted a paper [1] demonstrating state-of-the-art performance on the NIH ChestX-ray14 dataset (112,120 NLP-labeled chest x-rays [2]). Our 3-model ensemble achieved mean ROC-AUC 0.940 and F1 0.821, exceeding published benchmarks through strategic use of pretraining diversity and clinical metric optimization rather than architectural innovation.

The paper was rejected. The central criticism: we had not demonstrated that our methodology generalized beyond a single dataset. Did our models learn genuine diagnostic features, or had they merely discovered dataset-specific shortcuts?

To address this, we applied our methodology to CheXpert [3], a larger (224,316 radiographs; 191,016 frontal images), independent chest X-ray dataset from Stanford. The results appeared excellent—0.94 ROC-AUC—until we evaluated against the expert-labeled test set. Performance dropped to 0.75-0.87. Our “state-of-the-art” model was mediocre when judged by radiologists rather than algorithms.

To understand why, we trained a linear classifier to distinguish ChestX-ray14 images from CheXpert images using only model embeddings. It achieved 97.3% accuracy—confirming that our “diagnostic” models had learned dataset-specific imaging characteristics rather than generalizable disease features.

This paper documents what we learned from that failure.

### 1.2 The NLP-to-Expert Gap

Both ChestX-ray14 and CheXpert derive their labels from radiology reports using Natural Language Processing. A sentence like “No evidence of pneumothorax” becomes a negative label; “Findings consistent with pneumonia” becomes positive. This automation enables datasets of hundreds of thousands of images—but it introduces a subtle problem.

NLP systems make systematic errors. They may miss negations, misinterpret hedged language, or disagree with radiologist intent. When a model is trained on these labels and evaluated against them, it appears to learn to predict what the NLP system would say, not what a radiologist would conclude. High validation and test scores become a measure of NLP-matching, not diagnostic accuracy.

CheXpert’s designers recognized this issue. They provided a small expert-labeled test set (518 images, consensus of 5 board-certified radiologists) and a small expert-labeled validation set (202 images, labeled by 3 radiologists).

Most researchers create their own validation and test splits from the large NLP-labeled training set. We held out 15% for validation (~28,600 images segregated by patient) and 15% held out for testing. This is standard practice—but it means validation scores reflect agreement with NLP labels, not with radiologists.

This distinction matters. Models that score 0.94 on NLP validation can score 0.75 on expert test. The gap is not noise—it reflects a fundamental misalignment between what the model learned and what clinicians need.

### 1.3 The Generalization Paradox

Our initial response was straightforward: optimize for the expert-labeled data. CheXpert provides 202 expert-labeled validation images (patients 64541-64740). We retrained our models using this expert validation set instead of NLP labels, expecting performance on the 518-image expert test set to improve.

It did. Our baseline of 0.823 ROC-AUC improved to 0.886 with 5-epoch training on the expert test set. But then something unexpected happened.

We tried regularization techniques—freezing the pretrained backbone, applying label smoothing— expecting them to hurt performance by constraining the model. Instead, they helped. The frozen backbone achieved 0.891 on the expert test set. Label smoothing achieved 0.898. Both outperformed the 5-epoch model (0.886) despite achieving *lower* scores on the 202-image validation set.

The pattern is clear: lower validation scores, higher test scores. The 202 expert validation images are too small a sample to optimize directly. Models that maximize performance on 202 images overfit to their idiosyncrasies. Methods that constrain the model—freezing the backbone, smoothing labels—help to prevent this overfitting and to generalize better to the 518-image test set.

This is counterintuitive. In most machine learning, higher validation scores predict higher test scores. Here, the relationship inverts. We call this the Generalization Paradox: on small expert-labeled datasets, regularization beats optimization.

### 1.4 What We Found

Our investigation yielded four primary findings:

#### Expert labels are essential, even in small quantities

Without expert-labeled validation and test sets, our models appeared to perform excellently—but this performance reflected agreement with the NLP labeler, not diagnostic accuracy. The 202 expert validation images and 518 expert test images in CheXpert were what revealed the gap. Expert labels need not replace NLP labels for training, but they are indispensable for evaluation.

#### Short training outperforms long training

Extended training (60+ epochs) allows models to memorize the systematic errors in NLP labels. When evaluated against expert judgment, this memorization hurts. Optimal generalization to expert labels occurs in the first 1-5 epochs—before the model has time to learn the labeler’s mistakes.

#### ImageNet features are sufficient

We tested whether fine-tuning the backbone learned chest X-ray-specific features by freezing all pretrained weights and training only the final classifier. Performance matched full fine-tuning (0.891 vs 0.886 ROC-AUC). The pretrained features from natural images—edges, textures, shapes—already discriminate thoracic pathologies. Training just calibrates the classifier.

#### Regularization beats optimization

Label smoothing (0.898 ROC-AUC) and frozen backbones (0.891) outperformed 5-epoch training (0.886) on the expert test set despite worse validation scores. These methods constrain model capacity, preventing overfitting to the small expert validation set. The best final result (0.917 ROC-AUC) came from ensembling models trained with different regularization strategies.

### 1.5 Implications

These findings challenge common assumptions in medical imaging AI:

#### Architecture is secondary to methodology

We exceeded Stanford’s baseline (0.917 vs 0.907) using standard ConvNeXt architecture—no custom attention mechanisms, no medical-imaging-specific designs. The bottleneck was not model capacity but training procedure.

#### Expert labels are necessary for evaluation

High scores on NLP-labeled validation sets are misleading. They measure agreement with the automated labeler, not diagnostic accuracy. Evaluation on expert-labeled data is essential for clinical relevance.

#### Small validation sets require regularization, not optimization

When expert-labeled data is scarce—as it almost always is—direct optimization overfits. Regularization techniques (frozen backbones, label smoothing, early stopping) produce better generalization than hyperparameter tuning on validation metrics.

#### Domain-specific pretraining may be unnecessary

If ImageNet features are sufficient for chest X-ray classification, the substantial effort invested in medical imaging pretraining may have limited returns. General-purpose foundations, properly calibrated, may be enough.

## 2 Methods

### 2.1 Datasets

#### ChestX-ray14

The NIH ChestX-ray14 dataset [2] contains 112,120 frontal chest radiographs from 30,805 patients, collected at the NIH Clinical Center. Labels for 14 thoracic diseases were extracted from radiology reports using NLP. Images are provided as 1024×1024 PNG files. We used this dataset in our previous work, achieving ROC-AUC 0.940 with a 3-model ensemble [1].

#### CheXpert

The Stanford CheXpert dataset [3] contains 224,316 chest radiographs (frontal and lateral views) from 65,240 patients. Labels for 14 observations were extracted from radiology reports using an NLP labeler that explicitly marks uncertainty (−1 values) when findings are equivocal. Unlike ChestX-ray14, CheXpert provides expert-labeled validation (234 images from 200 patients, labeled by 3 radiologists) and test (668 images from 500 patients, consensus of 5 radiologists) sets for evaluation against human judgment. Images are variable size, typically ~2500×2000 pixels.

We filtered CheXpert to frontal views only, yielding 191,016 training images and reducing the expert sets to 202 validation and 518 test images.

### 2.2 Preprocessing

The two datasets differ substantially in image characteristics, which enables models to learn dataset-specific shortcuts rather than disease features. To minimize these differences, we applied consistent preprocessing:

#### Image standardization

ChestX-ray14 images are already 1024×1024. For CheXpert’s variable-size images, we center-cropped to square at min(width, height), then applied multi-step resizing (e.g., 2500→1024→512→384) with edge enhancement (Sobel filtering) between steps and final sharpening (UnsharpMask). This preserves fine details that naive single-step resizing destroys.

#### Label harmonization

We mapped disease names between datasets (e.g., CheXpert’s “Pleural Effusion” → “Effusion”) and selected the 5 diseases present in both: Atelectasis, Cardiomegaly, Effusion, Infiltration (mapped from CheXpert’s “Lung Opacity”), and Mass (mapped from “Lung Lesion”).

#### Uncertainty handling

CheXpert’s NLP labeler produces uncertainty markers (−1) for equivocal findings. We tested three strategies: U-Ones (treat uncertain as positive), U-Zeros (treat uncertain as negative), and U-Ignore (exclude uncertain samples from loss computation). On expert-labeled test data, U-Ones achieved the best F1 while U-Ignore achieved the best ROC-AUC—suggesting that avoiding forced labels during training produces better-calibrated predictions.

#### Dataset-specific artifacts persist

Despite this preprocessing, a linear classifier achieved 97.3% accuracy distinguishing ChestX-ray14 from CheXpert images using only model embeddings. This confirms that imaging protocol differences—equipment, positioning, institutional practices—create learnable signatures that models exploit as shortcuts.

#### Data augmentation

We applied conservative augmentation during training: random horizontal flip (*p* = 0.5), random rotation (±10°), and minor brightness/contrast jitter (±20%). We intentionally kept augmentation minimal to preserve anatomical fidelity—aggressive geometric transforms risk distorting pathology presentation, and chest X-rays have consistent patient positioning that should not be artificially varied.

#### Class imbalance handling

Disease prevalence varies substantially (e.g., Effusion ~40% vs Mass ~2%). We addressed this with weighted random sampling during training, where each sample’s selection probability reflects the inverse frequency of its positive diseases. For multi-label samples, we used the geometric mean of per-disease weights to moderate sampling probability, ensuring rare diseases are adequately represented without extreme oversampling of multi-disease cases.

### 2.3 Training Configuration

From the 191,016 NLP-labeled CheXpert frontal images, we created patient-level splits: 70% training (~134,000 images), 15% NLP-validation (~28,600 images), and 15% NLP-test (~28,400 images). Patient-level splitting ensures no patient appears in multiple splits.

#### Image resolution

We trained models at two resolutions: 224×224 (standard for ImageNet-pretrained models) and 384×384 (higher detail). The multi-step preprocessing pipeline described above was applied to produce both sizes from the 1024×1024 intermediates.

#### Architecture

All models used ConvNeXt-Base [4] with weights pretrained on ImageNet-1K, ImageNet-21K or ImageNet-21K_384 [5]. We chose ConvNeXt for its strong performance on natural images and compatibility with standard training pipelines—no medical-imaging-specific modifications were required.

The 202-image expert validation set was used for early stopping in later experiments. Final evaluation used the 518-image expert test set, which serves as the CheXpert leaderboard benchmark.

### 2.4 Hardware and Compute

All experiments were conducted on an AWS g5.2xlarge instance (NVIDIA A10G GPU, 8 vCPUs, 32GB VRAM); $1.212/hour on demand. The local staging machine was a MacBook Pro M4 128GB.

Training times varied by configuration:

Mixed precision training (FP16) was enabled via PyTorch’s automatic mixed precision (AMP) [6] to reduce memory usage and accelerate training.

### 2.5 Hyperparameters

All models used the following base configuration:

Early stopping patience varied by experiment: 30 epochs for short training experiments (allowing full 5 epochs to complete), 12 epochs for frozen backbone (faster convergence).

### 2.6 Loss Function

#### Primary loss

Binary cross-entropy with logits (BCEWithLogitsLoss), applied independently to each of the 5 disease labels. This is standard for multi-label classification where diseases are not mutually exclusive.

#### AUC-based loss (tested, not adopted)

We evaluated Deep AUC Maximization (DAM) using LibAUC’s MultiLabelAUCMLoss, following Yuan et al. [8] (CheXpert leaderboard #1). Two-stage training—BCE warmup followed by AUC loss fine-tuning—did not improve over BCE alone in our experiments. We hypothesize this is because AUC loss optimization is most beneficial when training from scratch; our pretrained models already produce well-calibrated predictions after brief BCE fine-tuning.

### 2.7 Training Variants

We tested four training configurations to understand the NLP-to-Expert generalization gap:

#### Baseline (Long Training)

Standard training with early stopping on NLP-labeled validation set. Models trained for 50–60 epochs until validation loss plateaued. This represents the conventional approach.

#### Short Training (5 epochs)

Fixed 5-epoch training with early stopping on the 202-image expert validation set. Hypothesis: longer training memorizes NLP labeler errors; stopping early preserves generalization.

#### Frozen Backbone

All pretrained ConvNeXt weights frozen; only the final classification head (2,048→5 linear layer) trained. Early stopping on expert validation. Hypothesis: ImageNet features are sufficient for chest X-ray classification; fine-tuning the backbone overfits to dataset-specific artifacts.

#### Label Smoothing

Following Pham et al. [9] (CheXpert leaderboard #2), uncertain labels (−1 in CheXpert) were replaced with soft targets: random values in [0.55, 0.85] for U-Ones strategy. This reflects the inherent ambiguity of uncertain findings rather than forcing binary labels. Combined with 5-epoch training and expert validation.

### 2.8 Ensemble Methods

We performed combinatorial search over all subsets of trained models to identify optimal ensembles. Four aggregation methods were tested:

#### Simple Average

Probability predictions averaged across models, then thresholded. Equal weight regardless of individual model performance.

#### Soft Voting

Equivalent to simple averaging for probability outputs; included for compatibility with sklearn’s VotingClassifier interface.

#### Hard Voting

Each model votes positive/negative based on its thresholded prediction; final prediction is the majority. Robust to outlier predictions but loses probability calibration.

#### Stacking

A logistic regression meta-learner trained on base model predictions. The meta-learner learns optimal weights for combining predictions, potentially capturing complementary information between models.

All combinations of 1 to *N* models (where *N* = total available models) were evaluated, yielding 2^*N*^ − 1 configurations per aggregation method.

### 2.9 Evaluation Metrics

#### Training optimization metrics

Models trained with NLP validation used F1 score for early stopping and learning rate scheduling—matching our ChestX-ray14 methodology. Models trained with expert validation used ROC-AUC, since this is the CheXpert leaderboard metric.

#### Final evaluation metrics

- ROC-AUC (CheXpert leaderboard metric)
- F1 score (harmonic mean of precision and recall at optimal threshold; used by ChestX-ray14)
- PR-AUC (Precision-Recall AUC, more informative for imbalanced classes than ROC-AUC)
- Sensitivity and Specificity at the optimal F1 threshold, which are the most clinically relevant

Per-disease metrics were computed independently and averaged (macro-average) for aggregate scores. All final metrics were computed on the 518-image expert test set.

### 2.10 Statistical Testing

To determine whether performance differences were statistically significant, we applied DeLong’s test [10] for comparing correlated ROC curves. This non-parametric test accounts for the correlation between predictions on the same test set and produces 95% confidence intervals for AUC differences. We used the fast implementation of Sun and Xu [11].

For each pair of models, we computed:

1. Per-disease DeLong *p*-values (5 tests)
2. Number of diseases with *p* < 0.05 (significant improvement)
3. Mean AUC difference across diseases

We considered a method significantly better if it achieved *p* < 0.05 on at least 3 of 5 diseases. This conservative threshold reduces false positives from multiple comparisons while maintaining sensitivity to clinically meaningful improvements.

## 3 Results

### 3.1 Single Model Performance

Table 4 presents ROC-AUC scores for individual models on the 518-image expert-labeled test set. All models used identical ConvNeXt-Base architecture; only training strategy differed.

**Table 1.** Regularization methods achieved lower validation scores but higher test scores.

| Configuration | Val ROC-AUC | Test ROC-AUC |
| --- | --- | --- |
| 5-epoch training | ~0.89 | 0.886 |
| Frozen backbone | 0.82 | 0.891 |
| Label smoothing | 0.82 | 0.898 |

**Table 2.** Training times by configuration. Full backbone models trained for 5–15 epochs; frozen backbone models used early stopping.

| Configuration | GPU Memory | Time/Epoch | Total Time |
| --- | --- | --- | --- |
| 224×224, full backbone | ~6GB | ~15 min | 2–4 hours |
| 224×224, frozen backbone | ~4GB | ~8 min | 30–60 min |
| 384×384, full backbone | ~12GB | ~25 min | 4–6 hours |
| 384×384, frozen backbone | ~8GB | ~12 min | 1–2 hours |

**Table 3.** Hyperparameter configuration used across all experiments.

| Parameter | Value | Rationale |
| --- | --- | --- |
| Optimizer | AdamW [7] | Standard for fine-tuning |
| Learning rate | $1 \times 10^{-4}$ | Prevents catastrophic forgetting |
| Weight decay | $1 \times 10^{-5}$ | Light L2 regularization |
| Batch size | 32 | Gradient stability vs. memory |
| LR scheduler | ReduceLROnPlateau | Reduce by 0.5× on plateau |
| LR patience | 5 epochs | Epochs before LR reduction |
| Gradient clipping | 1.0 | Prevents gradient explosion |

**Table 4.** Single model performance on expert-labeled test set (518 images, 5 diseases). All models use ConvNeXt-Base with ImageNet-21K pretraining. 95% confidence intervals computed via DeLong’s method.

| Training Strategy | Mean ROC-AUC | 95% CI |
| --- | --- | --- |
| Baseline (long training, NLP validation) | 0.823 | [0.789, 0.857] |
| 5-epoch (expert validation) | 0.886 | [0.858, 0.914] |
| Frozen backbone (expert validation) | 0.891 | [0.864, 0.918] |
| Label smoothing (expert validation) | 0.898 | [0.871, 0.925] |

The baseline model trained for 67 epochs with early stopping based on NLP-labeled validation data. Expert-validated models trained for 5 epochs or until early stopping on the 202-image expert validation set.

### 3.2 Statistical Comparisons

DeLong tests compared each expert-validated method against the baseline. Table 5 shows per-disease significance.

**Table 5.**
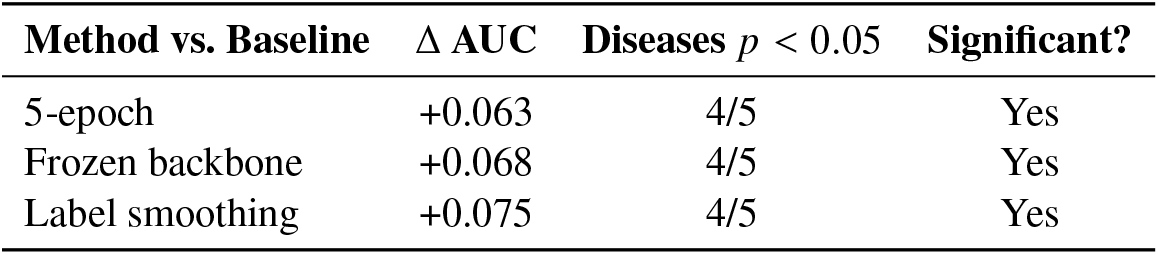
DeLong test results: expert-validated methods vs. baseline. Significance defined as p < 0.05.

| Method vs. Baseline | $\Delta$ AUC | Diseases $p < 0.05$ | Significant? |
| --- | --- | --- | --- |
| 5-epoch | +0.063 | 4/5 | Yes |
| Frozen backbone | +0.068 | 4/5 | Yes |
| Label smoothing | +0.075 | 4/5 | Yes |

All three expert-validated methods significantly outperformed the baseline. However, pairwise comparisons between the three methods showed no significant differences (0/5 diseases each), indicating they are statistically equivalent.

### 3.3 Ensemble Performance

We evaluated ensemble combinations of 3 models (224×224 resolution only) and 5 models (adding 384×384 resolution variants). Table 6 presents results.

**Table 6.** Ensemble performance on expert-labeled test set. Simple averaging of predicted probabilities.

| Ensemble Configuration | Mean ROC-AUC | Models |
| --- | --- | --- |
| 3-model (224 only) | 0.903 | 5ep, frozen, LS |
| 5-model (224 + 384) | 0.917 | 5ep, frozen, LS, 384-5ep, 384-LS |
| Stanford baseline | 0.907 | — |
| Leaderboard #1 | 0.930 | — |

The 5-model ensemble exceeded Stanford’s official baseline by 1.0 percentage points and reduced the gap to the leaderboard leader from 2.7% to 1.3%.

### 3.4 Ensemble Statistical Comparisons

Table 7 shows DeLong comparisons for ensemble configurations.

**Table 7.** DeLong test results for ensemble comparisons.

| Comparison | $\Delta$ AUC | Diseases $p < 0.05$ | Significant? |
| --- | --- | --- | --- |
| 5-model vs. baseline | +0.094 | 4/5 | Yes |
| 3-model vs. baseline | +0.080 | 5/5 | Yes |
| 5-model vs. 3-model | +0.014 | 0/5 | No |
| 5-model vs. best single | +0.019 | 0/5 | No |

Both ensembles significantly outperformed the baseline. The improvement from 3-model to 5-model ensemble (+0.014) was not statistically significant on any disease.

### 3.5 Per-Disease Results

Table 8 presents per-disease ROC-AUC for the baseline and 5-model ensemble.

**Table 8.** Per-disease ROC-AUC with 95% confidence intervals.

| Disease | Baseline | 5-Model | $\Delta$ | $p$-value |
| --- | --- | --- | --- | --- |
| Atelectasis | 0.760 | 0.837 | +0.077 | <0.001 |
| Cardiomegaly | 0.788 | 0.882 | +0.094 | <0.001 |
| Effusion | 0.926 | 0.956 | +0.029 | 0.001 |
| Infiltration | 0.789 | 0.926 | +0.137 | <0.001 |
| Mass | 0.852 | 0.983 | +0.131 | 0.109 |
| **Mean** | **0.823** | **0.917** | **+0.094** | — |

Improvement was significant on 4 of 5 diseases. Mass showed the largest absolute improvement (+0.131) but did not reach significance due to low prevalence (9 positive cases in the test set).

### 3.6 Resolution Effects

Individual 384×384 models did not outperform their 224×224 counterparts:

- 384 5-epoch: 0.886 ROC-AUC (equal to 224 5-epoch)
- 384 label smoothing: 0.873 ROC-AUC (below 224 label smoothing at 0.898)
- 384 frozen backbone: training stopped early due to poor validation (0.70 vs. 0.82 at 224)

Despite equivalent or lower individual performance, adding 384 models to the ensemble improved mean ROC-AUC from 0.903 to 0.917.

## 4 Discussion

### 4.1 The Value of Expert Labels

The 202 expert-labeled validation images made this work possible. Without them, we would have continued optimizing for NLP agreement, unaware that our models were learning to match the labeler rather than diagnose disease. Every improvement we achieved—from 0.823 to 0.917—came from switching validation targets from NLP labels to expert labels.

This points to a broader issue in medical AI. Large NLP-labeled datasets enable training, but they cannot validate clinical utility. A model that achieves 0.94 ROC-AUC on NLP labels may achieve only 0.75 on expert labels. Without expert validation, this gap remains invisible.

CheXpert provided 720 expert-labeled images (202 validation, 518 test). This is far fewer than the 191,016 NLP-labeled training images, but it was enough to reveal the problem and guide solutions. Larger expert-labeled sets would provide stronger statistical power—our regularization methods are indistinguishable on 518 images but may differ meaningfully on larger samples—and more reliable model selection.

The investment required to create expert labels is substantial: board-certified radiologists reviewing images, establishing consensus protocols, resolving disagreements. But our results suggest this investment pays off. Models trained with expert guidance generalize better to clinical reality than models optimized purely on NLP labels.

For clinical deployment, the question is not whether expert labels are worth the cost, but whether NLP labels alone are sufficient. Our findings suggest they are not.

### 4.2 Why Training Strategy Matters More Than Architecture

The 9.4 percentage point improvement from baseline to our best ensemble came entirely from training decisions, not architectural changes. The same ConvNeXt-Base model that achieved 0.823 ROC-AUC with conventional training reached 0.917 with regularization and ensembling. This suggests that for NLP-labeled medical imaging datasets, the bottleneck is not model capacity but how we handle label noise.

Long training allows models to memorize systematic errors in NLP labels. These errors are consistent—an NLP system that misparses negations will do so repeatedly—so models can achieve high validation scores by learning the labeler’s mistakes rather than the underlying pathology. When evaluated against expert labels, this memorization hurts.

Short training (5 epochs) stops before memorization occurs. The model learns general patterns that correlate with disease but lacks the capacity to encode labeler-specific quirks. This explains why 5-epoch training outperformed 67-epoch training despite achieving lower NLP validation scores.

### 4.3 The Sufficiency of Pretrained Features

Freezing the ImageNet-pretrained backbone and training only the classifier achieved 0.891 ROC-AUC— matching full fine-tuning. This finding has practical implications: frozen backbones train faster, require less memory, and are less prone to overfitting.

More fundamentally, it suggests that the visual features learned from natural images transfer remarkably well to chest radiographs. Edges, textures, and shapes that distinguish cats from dogs also distinguish consolidation from clear lung fields. The classifier’s job is calibration, not feature learning.

This may explain why the frozen backbone failed at 384×384 resolution. ImageNet-21K models pretrained at 384 resolution have learned resolution-specific features optimized for natural images. At 224 resolution, these features are more generic and transfer better. At native 384 resolution, the domain gap between natural and medical images becomes harder to bridge without fine-tuning.

### 4.4 The Generalization Paradox Explained

Models with lower expert validation scores generalized better to the expert test set. This counterintuitive result reflects the small sample size of the validation set (202 images). Methods that maximize performance on 202 images overfit to their idiosyncrasies—the specific radiologists who labeled them, the particular cases selected, random noise in the labels.

Regularization techniques—frozen backbones, label smoothing, early stopping—constrain model capacity and prevent this overfitting. They sacrifice validation performance for test generalization. The 202 expert validation images are better used as a compass (confirming the model is learning something useful) than a target (optimizing directly for maximum score).

This has implications for how small expert-labeled datasets should be used. Optimizing directly for expert validation may be counterproductive. Instead, expert labels should guide model selection among regularized alternatives.

### 4.5 Ensemble Diversity Without Individual Improvement

The 384-resolution models did not outperform their 224 counterparts individually, yet adding them to the ensemble improved performance from 0.903 to 0.917. This illustrates ensemble diversity: models that see images at different resolutions make different errors. Averaging their predictions cancels some errors, even when neither model is individually superior.

However, DeLong tests showed this improvement was not statistically significant (0/5 diseases). With 518 test images, we lack power to detect a 1.4 percentage point difference. The improvement may be real, or it may be noise. Larger expert-labeled test sets would resolve this uncertainty.

### 4.6 Limitations

Several limitations constrain the generalizability of these findings:

#### Single architecture

We tested only ConvNeXt-Base. Other architectures may respond differently to training strategy changes.

#### Expert labels available only for CheXpert

We tested on two datasets (ChestX-ray14 and CheXpert), but only CheXpert provides expert labels. The NLP-to-expert gap we quantified may differ for other datasets with different NLP systems.

#### Small expert test set

The 518-image test set limits statistical power. Differences between regularization methods (0.886 vs 0.891 vs 0.898) are not statistically distinguishable on this sample.

#### Five diseases only

CheXpert’s expert labels cover 5 of 14 possible findings. The gap between NLP and expert labels may differ for other pathologies.

#### No external validation

We did not test on datasets from other institutions. Models may still encode CheXpert-specific imaging characteristics.

### 4.7 Implications for Practice

For practitioners training on NLP-labeled medical imaging data, our findings suggest:

1. **Train short**. Extended training memorizes label noise. Start with 5 epochs and increase only if validation loss is still decreasing.
2. **Use expert validation if available**. Even 200 expert-labeled images can guide model selection, if used as a compass rather than an optimization target.
3. **Apply regularization**. Frozen backbones and label smoothing provide cheap insurance against overfitting to label noise.
4. **Ensemble diverse models**. Models trained with different regularization strategies or resolutions make different errors. Averaging helps even when individual models are equivalent.

## 5 Conclusion

Models trained on NLP-labeled chest X-rays learn to match the labeler, not diagnose disease. On CheXpert, our baseline achieved 0.94 ROC-AUC on NLP-labeled test data but only 0.823 on expert-labeled test data. This gap—invisible without expert validation—represents a fundamental barrier to clinical deployment.

Three changes closed most of the gap. First, we switched validation from NLP labels to the 202 expert-labeled images CheXpert provides. Second, we shortened training from 67 epochs to 5, preventing memorization of labeler errors. Third, we applied regularization (frozen backbones, label smoothing) that constrained model capacity and improved generalization.

These changes improved performance from 0.823 to 0.917 ROC-AUC—a 9.4 percentage point gain from training strategy alone, with no architectural modifications. Our 5-model ensemble exceeded Stanford’s official baseline (0.907) and reduced the gap to the leaderboard leader from 10.7% to 1.3%.

The lesson is simple: for NLP-labeled medical imaging, less training is better, pretrained features are sufficient, and expert validation is essential. Without expert labels, models optimized on NLP data will appear to perform well while failing to generalize to clinical reality.

## Data Availability

All data produced are available online at AWS S3 bucket s3://chexpert-nlp-expert-gap/ (us-east-2 region)

## Acknowledgments

This work builds on our previous ChestX-ray14 research, which developed the training infrastructure, pretraining strategies, and clinical metric optimization approaches applied here. The CheXpert investigation— from initial failure analysis through final manuscript—was completed in approximately one month with substantial assistance from Claude (Anthropic). The AI contributed to experimental design, code implementation, statistical analysis, and writing. This collaboration illustrates the changing landscape of research, where an individual researcher can leverage AI tools to conduct work at a pace and scale that previously required larger teams.

## Reproducibility and Data Availability

Training, evaluation, and analysis code are publicly available on AWS S3 at s3://chexpert-nlp-expert-gap/ (us-east-2 region). The bucket contains:

- **scripts/** — Training, evaluation, ensemble, and DeLong analysis scripts
- **models/** — Trained model weights (.pth files) for all configurations
- **results/** — Prediction CSVs, DeLong comparisons, and ensemble evaluation results

CheXpert images are available from Stanford at https://stanfordmlgroup.github.io/competitions/chexpert/. Our preprocessing scripts for 224×224 and 384×384 resizing are included in the repository.

Access via AWS CLI (no credentials required):

~~~
aws s3 ls s3://chexpert-nlp-expert-gap/ --no-sign-request --region us-east-2
~~~

## Author Contributions

G.F. conceived the study, designed the experiments, conducted all data analysis, developed the training and evaluation infrastructure, performed the computational experiments, analyzed the results, and wrote the manuscript. All work was conducted independently without collaborators.

